# The relationship between Lipoprotein A and other lipids with prostate cancer risk: A multivariable Mendelian randomisation study

**DOI:** 10.1101/2021.07.01.21259705

**Authors:** Anna Ioannidou, Eleanor L Watts, Aurora Perez-Cornago, Elizabeth A Platz, Ian G Mills, Timothy J Key, Ruth C Travis, The PRACTICAL consortium, CRUK, BPC3, CAPS, PEGASUS, Konstantinos K Tsilidis, Verena Zuber

**Affiliations:** Department of Epidemiology and Biostatistics, School of Public Health, Imperial College London, London, UK; Cancer Epidemiology Unit, Nuffield Department of Population Health, University of Oxford, Oxford, UK; Department of Epidemiology, Johns Hopkins Bloomberg School of Public Health, Baltimore, Maryland, USA; Sidney Kimmel Comprehensive Cancer Center at Johns Hopkins, Baltimore, Maryland, USA; Department of Urology and the James Buchanan Brady Urological Institute, Johns Hopkins University School of Medicine, Baltimore, Maryland, USA; Nuffield Department of Surgical Sciences, University of Oxford, Oxford, UK; Patrick G Johnston Centre for Cancer Research (PGJCCR), Queen’s University Belfast, Belfast, UK; Centre for Cancer Biomarkers (CCBIO), University of Bergen, 5021 Bergen, Norway; Department of Hygiene and Epidemiology, University of Ioannina School of Medicine, Ioannina, Greece

## Abstract

**Background:** Numerous epidemiological studies have investigated the role of blood lipids in prostate cancer (PCa) risk though findings remain inconclusive to date. The ongoing research has mainly involved observational studies which are often prone to confounding. This study aimed to identify the relationship between genetically predicted blood lipid concentrations and PCa.

**Methods and Findings:** Data for low-density lipoprotein (LDL) cholesterol, high-density lipoprotein (HDL) cholesterol, triglycerides (TG), apolipoprotein A (apoA) and B (apoB), lipoprotein A (Lp(a)) and PCa were acquired from genome-wide association studies in UK Biobank and the PRACTICAL consortium, respectively. We used a two-sample Mendelian randomisation (MR) approach with both univariable and multivariable (MVMR) models and utilised a variety of robust methods and sensitivity analyses to assess the possibility of MR assumptions violation. No association was observed between genetically predicted concentrations of HDL, TG, apoA and apoB and PCa risk. Genetically predicted LDL concentration was positively associated with total PCa in the univariable analysis but adjustment for HDL, TG and Lp(a) led to a null association. Genetically predicted concentration of Lp(a) was associated with higher total PCa risk in the univariable (OR_weighted median_ per sd = 1.091; 95% CI 1.028-1.157; P=0.004) and MVMR analyses after adjustment for the other lipid traits (OR_IVW_ per sd = 1.068; 95% CI 1.005-1.134; P = 0.034). Genetically predicted Lp(a) was also associated with advanced (MVMR OR_IVW_ per sd = 1.078; 95% CI 0.999-1.163; P=0.055) and early age onset PCa (MVMR OR_IVW_ per sd = 1.150; 95% CI 1.015,1.303; P = 0.028). Although multiple estimation methods were utilized to minimize the effect of pleiotropic traits, the presence of any unmeasured pleiotropy cannot be excluded and may limit our findings.

**Conclusions:** We observed that genetically predicted Lp(a) concentrations are associated with an increased PCa risk. Future studies are required to understand the underlying biological pathways of this finding, as it may inform PCa prevention through Lp(a)-lowering strategies.

## Introduction

Prostate cancer (PCa) is one of the most frequently diagnosed cancers in men [1] with 1,276,106 incident cases reported globally during 2018 [2]. There is high geographical heterogeneity of PCa incidence, which is reflected in a 40-fold difference in the age-adjusted incidence rates across the globe [3]. Several studies have argued that this could be attributed to the increased number of diagnoses in countries where the prostate specific antigen (PSA) screening is prevalent. Nevertheless, the basis for this heterogeneity remains poorly understood [1].

Given that PCa is also clinically heterogeneous, risk factors identified to date differ by disease aggressiveness [4]. In particular, established risk factors for total PCa are mainly non-modifiable, including older age, African-descent and genetics [5], whereas some potential risk factors for aggressive PCa include smoking, obesity [6], lower vitamin D and higher blood lipid levels [4] which are modifiable. Lipid-lowering therapies are cheap and well-established for lowering cardiovascular risk. Yet, there is no conclusive evidence that repurposed lipid-lowering drugs are effective for the prevention of PCa. It is therefore important to determine whether blood lipids increase PCa risk, especially lethal disease [7]. A meta-analysis of 14 prospective studies published in 2015 [10] did not observe significant associations between TC, HDL or LDL concentrations and risk of total or high-grade PCa, but high between-study heterogeneity was evident for most associations. Two meta-analyses have examined the role of statin use in PCa risk, and both observed inverse associations of statins and advanced PCa risk [11][12]. Nonetheless, whether these associations can be attributed to lower cholesterol itself or some other mechanism is unknown.

Observational studies may suffer from unobserved confounding and reverse-causation [13], which could explain inconsistent findings among studies. Mendelian randomisation (MR) uses genetic variants as proxies for the exposures of interest and, if carefully conducted, can complement observational research [14] and support triangulation of evidence. That is because genetic variants are randomly allocated to offspring by parents and independently assorted during meiosis, which minimise issues with reverse-causation and confounding [13,15]. In addition, most studies on lipids and PCa measure lipid levels only once which can lead to measurement error in the findings, whereas genetically predicted lipid levels capture lifelong expected levels. Previous MR research is limited to two studies that examined the role of HDL, LDL and TG in PCa risk overall and by disease stage and grade, and both reported null associations [16,17]. However, neither study adjusted for multiple lipid traits, which may have limited their findings given that different lipids are correlated and pleiotropic [18]. In this paper, we aim to identify whether genetically predicted lipid traits are associated with overall PCa risk, and in particular advanced and early age onset disease. We incorporated a univariable and multivariable MR (MVMR) framework to adjust for pleiotropic lipid effects, and examined the role of HDL, LDL and TG, as well as additional lipid-traits that have been underexamined to date, such as lipoprotein A (Lp(a)), apolipoprotein A (apoA) and apolipoprotein B (apoB).

## Methods

### Study populations

#### Blood Lipids Data

Genome-wide association (GWA) data for HDL, LDL, TG, Lp(a), apoA and apoB were available from UK Biobank, with information on over 13.7 million single-nucleotide polymorphisms (SNPs) [19]. Model adjustments in this UK Biobank GWAS included age, age^2, inferred_sex, interaction terms for age*inferred_sex and for age^2*inferred_sex, and the first 20 principal components. All measured serum biomarkers were approximately normally distributed except Lp(a), which was positively skewed. For consistency purposes, inverse rank-normalised data were used for all biomarkers. The heritability estimation from summary statistics (HESS) was applied in each of these lipids in a GWAS of Sinnott-Armstrong et al. [20] and heritability was reported as follows; HDL 36%, LDL 29%, TG 29%, Lp(a) 24%, apoA 31% and apoB 32% indicating strong genetic regulation of all lipid traits considered as exposures (Table S1). For the purpose of this research and to match with the PCa GWAS, only European ancestry male participants were included (N=167,020)

#### PCa Data

Summary association statistics for PCa risk were acquired from the PRACTICAL consortium and are based on Schumacher et al. [21]. The genotyping was performed using a custom array, namely the OncoArray. A two-stage imputation was performed, using the SHAPEIT20 and IMPUTEv221. For our analysis, we used total, advanced (metastatic or Gleason score (GS) >=8 or PSA>100 ng/mL or PCa death) and early age onset (PCa age <= 55) PCa. Study participants for total PCa make up to a total of 79,166 cases and 61,106 controls, advanced PCa cases include 15,167 participants and 58,308 controls, whereas early age onset PCa includes 6,988 cases and 44,256 controls. All participants were of European ancestry.

#### Assumptions

The following assumptions were made for all our MR analyses and are described in combination for both the univariable and MVMR approaches [22].

1. ***Relevance:*** Genetic variants are associated with the exposure of interest in the case of univariable MR, whereas for MVMR, they are associated with at least one of the exposures.
2. ***Exchangeability:*** Genetic variants are independent of all confounders of the exposure-outcome association for the univariable MR, whereas in the MVMR, variants are independent of all confounders of each of the exposure-outcome associations.
3. ***Exclusion-restriction:*** Genetic variants are independent of the outcome given the exposure/es and all the confounders.

### Main MR analyses

All MR analyses were performed in R version 4.0.0. Due to the availability of exposure (blood lipids) and outcome (PCa) data from two different sources, we used a two-sample MR study design. In the univariable MR, SNPs that satisfied genome-wide significance (*P< 5 ×* 10^−8^) were selected for each trait. After removing any inconsistencies with respect to the effect and non-effect alleles, we harmonised the data so that the exposure and outcome datasets would have the same effect allele. We used the TwoSampleMR package version 0.5.4 to clump the data using a threshold of *r2<0*.*001* to identify and remove any SNPs in high linkage disequilibrium (LD). All SNPs left after clumping were considered as the instrumental variables (IVs). We firstly ran the univariable analysis on all blood lipids for both total and advanced PCa, whilst for the early age onset PCa we focused our univariable analysis primarily on Lp(a). For the main estimation methods of the univariable analyses, we performed the inverse variance weighting (IVW) [23,24] and weighed median [13] approaches, and we additionally applied the MR-Egger [25] approach.

To adjust for different lipid traits in our models, we performed a MVMR analysis. We chose to exclude apoA and apoB to avoid multicollinearity issues due to their high correlation with HDL and LDL, respectively (*r*_*apoA,HDL*_ *=0*.*978; P <2*.*2×*10^−6^*/r*_*apoB,LDL*_ *=0*.*984; P<2. 2×*10^−6^). The minimum *p*-value across the remaining lipids was computed and selection of SNPs was based on those that satisfied genome-wide significance through the minimum *p-*value (*P<5×*10^−8^). After harmonisation was performed, we clumped the data based on a threshold of *r2<0*.*001*. The main estimation method performed was the IVW, while we additionally implemented the MR-Egger estimate [26] to control for any remaining unmeasured pleiotropy.

### Sensitivity MR analyses

As we observed a positive finding for Lp(a) and PCa outcomes, we performed the following sensitivity analyses in our univariable MR considering only Lp(a) as exposure for total, advanced and early age onset of PCa.

1. ***Sensitivity analysis 1:*** As an attempt to increase the statistical power of the univariable MR, we used an eased clumping threshold of *r2<0*.*01* and re-fitted the models based on a larger set of IVs.
2. ***Sensitivity analysis 2:*** Variants that were used as IVs for Lp(a) in the Burgess et al. paper [27] based on a clumping threshold of *r2<0*.*4*, were separately fitted to the univariable models to validate findings on a different IV set. 43 IVs were initially proposed in the paper, but only 28 of those were available in our current datasets and satisfied genome-wide significance. The univariable models were re-fitted based on these 28 IVs.
3. ***Sensitivity analysis 3:*** As the *LPA* gene (chromosome 6: 160,531,482-160,664,275) is the main gene associated with Lp(a) concentrations and explains about 70-90% of its variability [28], we selected variants located in the *LPA* gene based on a clumping threshold of *r2<0*.*001* to represent strong biological instruments and potentially support the effect of Lp(a). Four such variants were identified and subsequently utilised as IVs.
4. ***Sensitivity analysis 4:*** Additional robust estimation methods were utilised as part of our sensitivity analyses to control and/or test for horizontal pleiotropy. These included the MR-PRESSO [29] and contamination mixture [30].

As obesity may be considered a probable confounder for lipids and PCa [16], we also performed an additional adjustment for body mass index (BMI) in all the MVMR models, using genetic association data for BMI from UK Biobank [19]. Lp(a) is assembled in the liver [31], whereas liver function/disease has been proposed to influence PCa detection and outcomes [31-32]. Genetic associations for aspartate aminotransferase (AST) were thereby adjusted in a MVMR model including Lp(a) and total PCa. In addition, as kidney disease has been suggested to affect Lp(a) concentrations [34], and creatinine was previously associated with PCa risk [35], we performed another MVMR analysis using Lp(a), creatinine and total PCa to control for kidney function. Both genetic associations for AST and creatinine were acquired from UK Biobank [19]. Throughout our analyses, we considered significant estimates based on the 95% confidence level. We additionally estimated a Bonferroni-corrected p-value for the main univariable analyses on total and advanced PCa, to adjust for the multiple tests performed on each outcome. The total number of tests is reflected upon the number of different lipids we considered for each PCa outcome. Throughout the results section, nominally significant p-values are reported.

## Results

Descriptive statistics for lipid measurements were available from UK Biobank [19] and can be seen in S2 Table. Throughout this section we report results based solely on the IVW and weighted median methods. Results from the additional methods we used, including MR-Egger (Table S3-S6), MR-PRESSO and contamination mixture estimates (Table S3), and MVMR analyses adjusting for BMI, AST and creatinine (Tables S6-9), can be found in the supplement and were in general in agreement with the main analyses presented in the text below. The marginal associations of the genetic instruments with exposures, outcomes and confounders/mediators are shown in Tables S10-15.

### Univariable MR

#### Total PCa

The univariable MR analysis showed that genetically predicted HDL (*OR*_*IVW*_= 0.994; 95% CI =[0.942,1.051]; P=0.825), TG (*OR*_*IVW*_ =1.026; 95% CI = [0.961,1.105]; P=0.449), apoA (*OR*_*IVW*_ =1.025; 95% CI = [0.970,1.083]; P=0.372) and apoB (*OR*_*IVW*_ =1.026; 95% CI = 0.961,1.094]; P=0.411) concentrations were not associated with total PCa risk (Table S4). In contrast, the odds ratio of total PCa was 1.088 per standard deviation (sd) increase in genetically predicted LDL (95% CI = [1.010,1.162]; P=0.016). This association was however not supported by the weighted median approach (OR=1.016; 95% CI = [0.942,1.094]; P=0.669). This raised concerns for potential pleiotropic effects present in our model and results were further assessed in the multivariable model.

Genetically predicted Lp(a) had an insignificant association on total PCa as estimated from the IVW (*OR*_*IVW*_= 1.066; 95% CI = [0.909,1.249]; P=0.431) method (Table 1), but the odds ratio of total PCa in the weighted median approach was 1.091 per sd increase in genetically predicted Lp(a) (95% CI =[1.028,1.157]; P=0.004). Alteration of the clumping threshold in sensitivity analysis 1 resulted in a higher number of IVs fitted to our model, which supported a significant effect estimate for Lp(a) in both the IVW (*OR*_*weighted median*_ = 1.076; 95% CI = [1.016,1.114]; P=0.012) and weighted median approaches (*OR*_*weighted median*_ = 1.066; 95% CI = [1.012,1.123]; P=0.016). Sensitivity analysis 2, which included IVs according to the Burgess et al. paper [27], also supported a relationship between genetically elevated Lp(a) and total PCa (0R =1.037; 95% CI = [1.009,1.066]; P=0.010, *OR*_*weighted median*_ = 1.044; 95% CI = [1.026,1.061]; P=6.58×10-7). Sensitivity analysis 3, which involved variants located in the *LPA* gene only, supported an even stronger OR (*OR*_*weighted median*_ =1.439; 95% CI = [1.280,1.619]; P=1.80×10^−9^) for total PCa per sd increase in genetically predicted Lp(a).

**Table 1.** Univariable estimates of genetically predicted Lp(a) on each PCa outcome. Results are based on the inverse variance weighting (IVW) and weighted median estimation methods. ORs for each PCa outcome are reported per sd increase in genetically predicted Lp(a). The main analysis included IVs based on a clumping threshold of 0.001, sensitivity analysis 1 is based on an eased clumping threshold of 0.01, sensitivity analysis 2 is based on a different IV set from another paper and finally sensitivity analysis 3 is based upon variants located in the *LPA* gene. Associations of P<0.05 are shown in bold. .P ∈ (0.05,0.1], * P ∈ (0.01,0.05], ** P ∈ (0.001,0.01], *** P ∈ (0,0.001]. † Padjusted is the Bonferroni corrected p-value, taking into account the total number of tests performed in the main analysis for total and advanced PCa. This reflects a total of 5 univariable analyses performed on each outcome (one for each lipid). For the early age onset of PCa only Lp(a) was fitted to the model and thus no correction was applied to the p-value.

|  |  | *Method* | *OR* | *95% CI* | *P* | *Padjusted†* |
| --- | --- | --- | --- | --- | --- | --- |
| *Total PCa* | **Main Analysis** | IVW | 1.066 | [0.909,1.249] | 0.431 | 1 |
|  |  | **Weighted Median** | **1.091** | **[1.028,1.157]** | **0.004\*\*** | **0.024\*** |
|  | **Sensitivity Analysis 1** | IVW | **1.076** | **[1.016,1.114]** | **0.012\*** | - |
|  |  | **Weighted Median** | **1.066** | **[1.012,1.123]** | **0.016\*** | - |
|  | **Sensitivity Analysis 2** | IVW | **1.037** | **[1.009,1.066]** | **0.010\*\*** | - |
|  |  | **Weighted Median** | **1.044** | **[1.026,1.061]** | **6.582x10<sup>-7</sup>\*\*\*** | - |
|  | **Sensitivity Analysis 3** | IVW | 1.228 | [0.960,1.570] | 0.104 | - |
|  |  | **Weighted Median** | **1.439** | **[1.280,1.619]** | **1.799x10<sup>-9</sup>\*\*\*** | - |
| *Advanced PCa* | **Main Analysis** | IVW | 1.064 | [0.910,1.245] | 0.435 | 1 |
|  |  | **Weighted Median** | 1.071 | [0.973,1.179] | 0.158 | 0.948 |
|  | **Sensitivity Analysis 1** | IVW | 1.051 | [0.981,1.127] | 0.158 | - |
|  |  | **Weighted Median** | 1.051 | [0.974,1.135] | 0.197 | - |
|  | **Sensitivity Analysis 2** | IVW | 1.024 | [0.992,1.058] | 0.139 | - |
|  |  | **Weighted Median** | **1.033** | **[1.001,1.065]** | **0.046\*** | - |
|  | **Sensitivity Analysis 3** | IVW | 1.226 | [0.957,1.570] | 0.107 | - |
|  |  | **Weighted Median** | **1.388** | **[1.213,1.590]** | **2.138x10<sup>-6</sup>\*\*\*** | - |
| *Early age onset PCa* | **Main Analysis** | IVW | 1.169 | [0.927,1.473] | 0.188 | - |
|  |  | **Weighted Median** | **1.257** | **[1.107,1.426]** | **4.000x10<sup>-4</sup>\*\*\*** | - |
|  | **Sensitivity Analysis 1** | IVW | **1.215** | **[1.096,1.349]** | **2.028x10<sup>-4</sup>\*\*\*** | - |
|  |  | **Weighted Median** | **1.217** | **[1.084,1.365]** | **0.001\*\*\*** | - |
|  | **Sensitivity Analysis 2** | IVW | **1.076** | **[1.027,1.126]** | **0.002\*\*** | - |
|  |  | **Weighted Median** | **1.079** | **[1.036,1.124]** | **3.201x10<sup>-4</sup>\*\*\*** | - |
|  | **Sensitivity Analysis 3** | IVW | 1.481 | [0.907,2.418] | 0.116 | - |
|  |  | **Weighted Median** | **1.502** | **[1.276,1.770]** | **1.038x10<sup>-6</sup>\*\*\*** | - |

#### Advanced PCa

The univariable MR analysis did not reveal any significant association between blood lipids and advanced PCa risk (HDL; *OR*_*IVM*_ = 0.977; 95% CI = [0.905,1.051]; P =0.552, LDL; *OR*_*IVM*_ =1.067; *OR*_*IVM*_ 95% CI = [0.970,1.74]]; P=0.191, TG; 0R = 1.004; 95% CI =[0.923,1.094]; P = 0.921, Lp(a) ; *OR*_*IVM*_ = 1.064; 95% CI = 0.910,1.245]; P = 0.435, ApoA; 0R = 1.001; 95% CI = [0.932,1.073],P = 0.991, ApoB; 0R = 0.992; 95% CI = [0.914,1.073], P = 0.837) (Table 1, Table S5). However, sensitivity analysis 2 for Lp(a), which included IVs of the Burgess et al. paper [27], supported an association between genetically elevated Lp(a) (*OR*_*weighted median*_ = 1.033; 95% CI= [1.001,1.065]; P=0.046) and advanced PCa. In addition, sensitivity analysis 3, which restricted to variants in the *LPA* gene, supported an association between genetically elevated Lp(a) and advanced PCa (*OR*_*weighted median*_ = 1.388; 95% CI = [1.213,1.590]; P=2.14×10^−6^).

#### Early age onset of PCa

Genetically predicted Lp(a) was associated with an increased risk of early age onset PCa in the main univariable analysis (*OR*_*weighted median*_ =1.257; 95% CI = [1.107,1.426]; P=4.00×10^−4^) (Table 1). All univariable sensitivity analyses performed, confirmed a significant relationship between genetically elevated Lp(a) and early age onset of PCa. [(Sensitivity analysis 1; *OR*_*IVM =*_1.215; 95% CI = [1.096, 1.349]; P=2.03×10^−4^, 0R = 1.217; 95% CI = [1.084,1.365]; P=0.001), (Sensitivity analysis 2; *OR*_*IVM*_= 1.076; 95% CI = [1.027,1.126]; p-value=0.002, *OR*_*weighted median*_ = 1.079; 95% CI =[1.036,1.124]; P=3.20×10-4), (Sensitivity analysis 3; *OR*_*weighted median*_ = 1.502; 95% CI = [1.276,1.770]; P=1.04×10^−6^)].

### Multivariable MR

As an attempt to control for pleiotropic pathways that could arise from the relationship between different lipid traits, we incorporated a MVMR model including Lp(a), HDL, LDL and TG jointly as exposures for each PCa outcome. The significant association observed between genetically predicted LDL and total PCa in univariable MR attenuated in the multivariable MR model and was no longer significant (OR = 1.052; 95% CI = [0.973,1.134]; P=0.183) (Table 2). However, after adjusting for HDL, LDL and TG, genetically predicted Lp(a) remained significantly and positively associated with total PCa (OR = 1.068; 95% CI = [1.005,1.134]; P=0.034). Additional adjustment for BMI led to an almost unaltered OR for total PCa risk per sd increase in genetically predicted Lp(a) (OR = 1.066; 95% CI = [1.008,1.129]; P=0.026) (Table S7). Genetically predicted Lp(a) was associated at borderline significance with advanced PCa after adjusting for multiple lipid traits (OR= 1.078; 95% CI = [0.999,1.163]; P=0.055). Additional adjustment for BMI led to an OR of 1.075 (95% CI: [1,1.155]; P=0.050). Genetically elevated Lp(a) remained significantly associated with early age onset of PCa (OR= 1.150; 95% CI = [1.015,1.303]; P = 0.028), in agreement with the univariable MR analysis. Adjustment for BMI yielded a similar effect of 1.155 (95% CI = [1.029,1.297]; P = 0.015). IVW estimates for all lipids from the MVMR can be seen in Table 2 below, whereas the IVW BMI-adjusted results can be found in Table S7. We compared the multivariable Lp(a) estimates from all the analyses performed on total, advanced and early age onset PCa with the univariable estimates and additional sensitivity analyses through a panel of three distinct forest plots (Fig 1). IVs according to variants in the *LPA* gene (Sensitivity Analysis 3), supported the strongest effect between genetically predicted Lp(a) concentrations and each PCa outcome.

**Table 2.** MVMR results for each PCa outcome. Each estimate (OR) is based on the multivariable IVW method and represents the direct effect of the risk factor on the respective outcome after controlling for the other three biomarkers in MVMR. ORs are reported per sd increase in the respective biomarker. Genetically elevated Lp(a) is significantly associated with total and early age onset PCa, whereas it is associated also at borderline significance with advanced PCa. Associations of P<0.05 are shown in bold. .P ∈ (0.05,0.1], * P ∈ (0.01,0.05], ** P ∈ (0.001,0.01], *** P ∈ (0,0.001].

|  | *Biomarker* | *OR* | *95% CI* | *P* |
| --- | --- | --- | --- | --- |
| **Total PCa** | HDL | 1.009 | [0.946,1.077] | 0.775 |
|  | LDL | 1.052 | [0.973,1.134] | 0.183 |
|  | TG | 1.026 | [0.953,1.106] | 0.485 |
|  | **Lp(a)** | **1.068** | **[1.005,1.134]** | **0.034\*** |
| **Advanced PCa** | HDL | 0.993 | [0.917,1.077] | 0.872 |
|  | LDL | 1.007 | [0.916,1.106] | 0.892 |
|  | TG | 1.003 | [0.914,1.101] | 0.952 |
|  | Lp(a) | 1.078 | [0.999,1.163] | 0.055. |
| **Early age onset PCa** | HDL | 1.024 | [0.894,1.174] | 0.732 |
|  | LDL | 1.112 | [0.948,1.305] | 0.192 |
|  | TG | 1.062 | [0.908,1.242] | 0.450 |
|  | **Lp(a)** | **1.150** | **[1.015,1.303]** | **0.028\*** |

**Fig 1.**
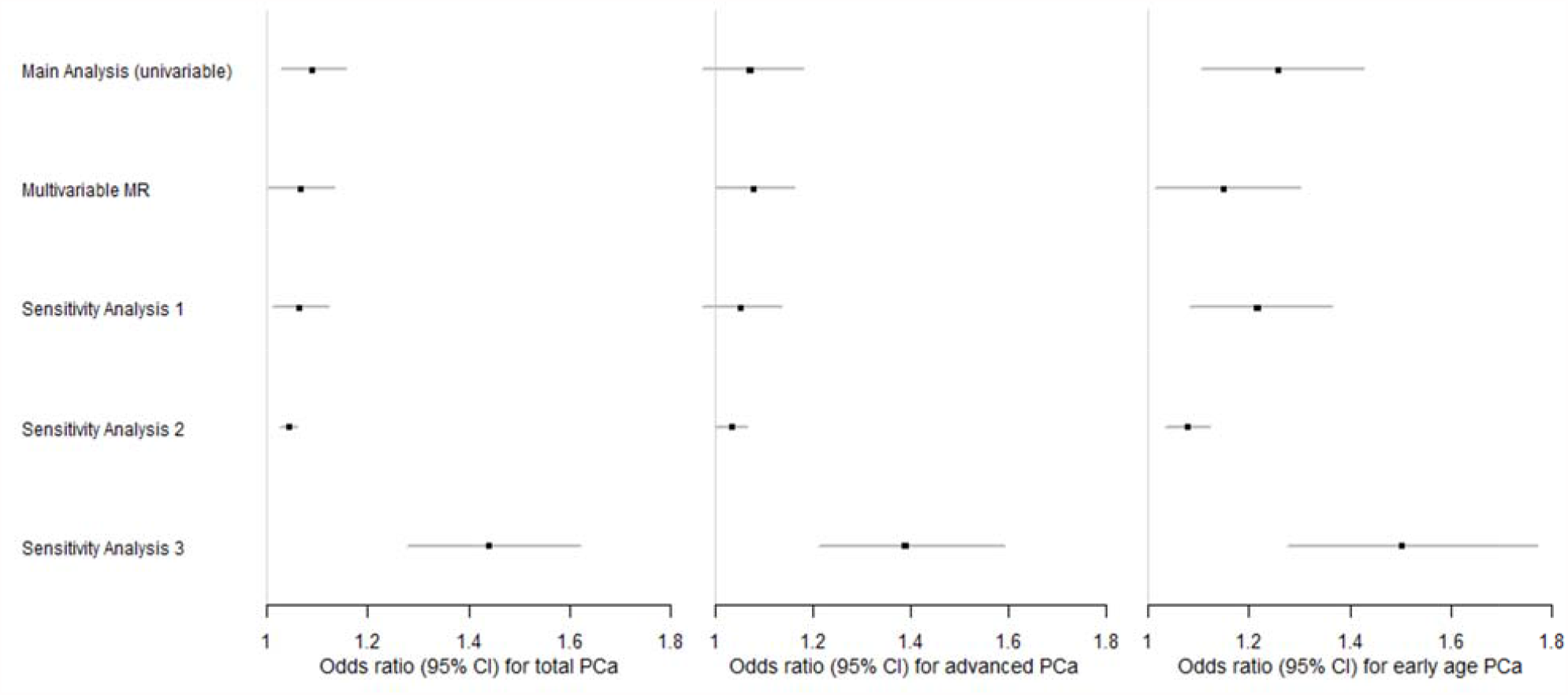
Forest plots of the Lp(a) effects observed in different analyses based on each PCa type. The main and sensitivity analyses estimates are based on the weighted median approach, whereas MVMR includes the IVW estimates. Sensitivity analyses 1-3 refer to the univariable models. Sensitivity analysis 1 is based on an eased clumping threshold of 0.01, sensitivity analysis 2 includes an IV set based on another paper and finally sensitivity analysis 3 is based upon variants located in the *LPA* gene. Each square represents the OR for each PCa outcome, reported per sd increase in the biomarker, with the 95% CI represented by the error bars.

## Discussion

Our MR analyses provide evidence that genetically predicted Lp(a) concentration is associated with risk of total and early age onset PCa and suggestive evidence for an association with advanced PCa. The majority of the different methods we used for our univariable analyses agreed for total and early age onset PCa. Lp(a) was only significantly associated with advanced PCa in two of the univariable sensitivity analyses. IVs located in the *LPA* gene supported the strongest and most significant Lp(a) associations for total, advanced and early age onset PCa. Given the strong regulation of Lp(a) levels by the *LPA* gene region [28], the latter findings are based on strong instruments with a clear biological function. Adjustment for multiple lipid traits and BMI in the MVMR models, further supported a significant association of genetically predicted Lp(a) on total, advanced and early age onset PCa.

The null associations observed for genetically predicted HDL on total PCa agree with findings from two previous MR analyses [16][17], and the null findings for TG are also supported in the Bull et al. [16] paper. In our analysis, there was some evidence for a significant LDL association with total PCa risk, though this was likely a false indication due to pleiotropy, as suggested by the MVMR model which indicated no association with LDL. As Lp(a) includes an LDL component [36], the attenuation of LDL to the null in the MVMR could be attributed to independent actions of Lp(a) itself, as we did not observe any association between other Lp(a) components and PCa risk. Alternative explanations are that Lp(a) concentrations are less affected by statins compared to LDL [37], thus genetically predicted Lp(a) may be more accurate for current actual levels than genetically predicted LDL, or that the association for Lp(a) dominates over LDL due to the high between-person variability of Lp(a) [20]. The authors of the Bull et al. paper [16] suggested a potential role of LDL and TG in advanced/high-grade PCa; our findings for LDL and TG in advanced PCa risk are not in agreement with theirs. Our analyses included adjustment for multiple lipid traits in contrast with the previously mentioned papers, which we believe plays a vital role in MR analysis modelling blood lipids, given the high correlation between them. As far as we are aware, no previous MR study has investigated the role of apoA and apoB in PCa risk. Our null results are nonetheless in agreement with observational studies by Katzke et al [8], which involved the prospective EPIC–Heidelberg cohort and Borgquist et al [38], which was based on the prospective Malmö Diet and Cancer Study (MDCS).

To the best of our knowledge, no previous MR study has examined the role of Lp(a) in PCa risk. The positive association observed for Lp(a) in total PCa was supported by the observational study of Katzke et al. [8]. Results showed that top vs. bottom quartile levels of Lp(a) were associated with a 47% higher risk of PCa (OR=1.47; 95% CI = 1.06-2.04). Wang F et al. [39], another observational study, examined the role of Lp(a) in high-risk PCa via a multivariable regression adjusted for age, BMI, hypertension, diabetes, coronary artery disease, and lipid-lowering drugs. They observed that high Lp(a) levels were positively associated (OR_Q4 vs. Q1_=2.687; 95% CI = 1.113-6.491; P = 0.028) with high-risk PCa, which agrees with our findings for advanced PCa in the MVMR analysis. In addition, a recent large prospective cohort among 211,754 men in UK Biobank [40] observed a suggestive positive association between Lp(a) and PCa risk (hazard ratio_per SD_ =1.02; 95% CI: [0.99,1.06]). Our literature review did not reveal any studies investigating the role of Lp(a) in early age onset PCa.

A range of different biological mechanisms have been proposed to explain pro-cancer effects of cholesterol at the cellular level, including cell proliferation, inflammation, membrane organization, and steroidogenesis [41]. It is unclear whether total cholesterol or any lipoprotein particle is the causal factor, and the potential pathophysiological mechanisms of Lp(a) have not been well-studied. However, emerging evidence from the cardiovascular literature supports pleiotropic functions of Lp(a) and complex mediation pathways with other lipid particles [42]. Lp(a) is highly heritable (heritability=24%) [20], with the majority of individuals having low Lp(a) levels. However, African Americans which are known to have the highest risk for PCa, tend to also have higher circulating Lp(a) levels [28]. It has been previously observed that mean Lp(a) concentrations for African Americans are 106 (60-180) nmol/l, whereas Caucasians such as non-Hispanic whites have mean Lp(a) concentrations of 24 (7.2-79.2) nmol/l [43]. Although the exact explanation behind ethnic discrepancies in PCa is currently unknown, it has been hypothesised that access to health care may play a partial role in this. Yet, given that disparities in PCa risk are apparent regardless of cancer detection issues, it is likely that biological factors are key drivers of this phenomenon [44]. Two recent papers have further provided evidence of a different immune response [45] and inflammatory signalling [46] for African Americans vs. Caucasians, which can be linked to their poorer PCa prognosis. Considering Lp(a) as a modifier of the immune/inflammatory response [47], the increased Lp(a) concentrations in African Americans and our observed association between genetically elevated Lp(a) and PCa, we hypothesise that Lp(a) may partially account for some of the observed discrepancies in PCa risk by ethnicity. Future large-scale genomic studies in African ancestry populations [48] would be required to evaluate the hypothesis that Lp(a) can explain discrepancies in PCa risk by race.

We note several limitations to our research. Currently, there is no direct way to prove that the second and third MR assumptions hold and as such, violations would result in biased MR estimates. A large number of robust methods and sensitivity analyses were used to probe into potential violations mainly due to horizontal pleiotropy, but its presence cannot be excluded. The samples analysed for our main MR analyses were restricted to Europeans to avoid issues with heterogeneity, which is required for a two-sample MR [49]. However, this may affect generalisability of the results, which are restricted to those of European ancestry. The number of variants associated with Lp(a) was limited in comparison to other lipids. Initially, 5894 variants were identified to be associated with Lp(a) at GWAS significance, whereas all other lipids had more than 10,000 associated variants. This then resulted in a final sample size of 10 variants due to LD clumping in the main univariable analysis, which may have decreased our statistical power. However, after relaxing the LD clumping threshold in our sensitivity analyses, we included more variants, the findings of which corroborated the main results. In addition, some previous observational studies have suggested potential threshold effects for cholesterol concentrations and PCa [50,51], which cannot be studied in two-sample MR with summary-level data, and future one-sample MR studies are warranted.

Apart from the caveats in our study, there are also several strengths that should be noted. We used an MR study design, in which the outcome of interest is compared between genotypes, analogous to that between treatment and placebo groups in a randomised controlled trial. However, inference should be made with great caution as alterations of genetically predicted risk factors are not identical to those due to a drug or dietary intervention [52]. Secondly, as lipids are dependent on each other for their main functionalities [18], it is important to control for pleiotropic pathways that may arise from these dependencies. One method to do so, is via the use of MVMR which allows to include genetic information on exposures that may correlate with each other into a joint multivariable model [53] and our study forms the first such MVMR conducted to investigate the relationship between various lipid traits and PCa risk. Thirdly, the use of UK Biobank data allowed us to include information on underexamined lipid traits such as Lp(a), apoA and apoB, in comparison to previous PCa studies which mainly considered HDL, LDL and TG. In addition, we have sex-specific genetic associations and this allowed us to work with male-specific data which are more relevant to PCa. Finally, our analyses are based on large sample sizes which were acquired from UK Biobank [19] and the PRACTICAL consortium [21].

In summary, findings from this study point towards a positive association between genetically predicted Lp(a) concentrations and risk of total, advanced and early age onset PCa. Screening for high Lp(a) concentrations could possibly be investigated in the future to identify high-risk groups for PCa. Given that Lp(a) concentrations depend significantly on genetics [54], modification of Lp(a) levels may be achieved by developing Lp(a)-lowering drugs [55] that might be on the horizon. A personalised approach in repurposing lipid drugs that target Lp(a) directly for high-risk individuals could consequently be considered, upon replication of our findings, to study their effectiveness against PCa prevention. The mechanisms behind the observed association remain however unclear given the uncertainty underlining the pleiotropic physiological functions of the *LPA* gene itself, which controls about 70-90% of the Lp(a) variability [36,54]. Further research into this complex gene such as colocalization analysis would be required to understand more of its functionality and consequently its role in PCa risk.

## Data Availability

Summary-level data on genetic associations with blood lipids in men (http://www.nealelab.is/uk-biobank) and overall prostate cancer (http://practical.icr.ac.uk/) are in the public domain. Summary-level data on genetic associations with subtypes of prostate cancer are available from the Practical Consortium.

http://practical.icr.ac.uk/

## Supporting information

**S1 Table. Heritability of each blood lipid as estimated in a GWAS of Sinnott-Armstrong et al [20]**. The estimates represent total heritability and the methodology followed was the Heritability estimation summary statistics (HESS). The number of SNPS fitted in each model for the main univariable analysis is also reported.

**S2 Table. Descriptive statistics of each blood lipid**. Lp(a) is positively skewed, whereas the rest of the lipids are approximately normally distributed. Measurements are based on all samples (both sexes) in UK Biobank [19].

**S3 Table. MR-Egger, MR-Presso and Contamination Mixture estimates for Lp(a) on each PCa outcome**. The contamination mixture method may indicate two distinct CIs associated with a single estimate. Associations of P<0.05 are shown in bold. .P ∈ (0.05,0.1], * P ∈ (0.01,0.05], ** P ∈ (0.001,0.01], *** P ∈ (0,0.001].

**S4 Table. Univariable MR estimates for each lipid on total PCa**. Genetically elevated LDL is significantly associated with total PCa only through the IVW approach, whereas the pleiotropy-robust methods do not support this association. Associations of P<0.05 are shown in bold. .P ∈ (0.05,0.1], * P ∈ (0.01,0.05], ** P ∈ (0.001,0.01], *** P ∈ (0,0.001].

**S5 Table. Univariable MR estimates for each lipid on advanced PCa**. None of these lipids are associated with advanced PCa.

**S6 Table. MR-Egger estimates for Lp(a) and each PCa outcome**. Sensitivity Analyses refer to the univariable MR and are as follows; Sensitivity Analysis 1 includes variants according to an eased clumping threshold of r2<0.01, sensitivity analysis 2 includes SNPs for Lp(a) according to a different IV set, whereas sensitivity analysis 3 is based on variants included in the LPA gene. Associations of P<0.05 are shown in bold. .P ∈ (0.05,0.1], * P ∈ (0.01,0.05], ** P ∈ (0.001,0.01], *** P ∈ (0,0.001].

**S7 Table. IVW estimates from the MVMR model adjusted for BMI**. Genetically elevated Lp(a) is associated with overall, advanced and early onset of PCa in these models. Associations of P<0.05 are shown in bold. .P ∈ (0.05,0.1], * P ∈ (0.01,0.05], ** P ∈ (0.001,0.01], *** P ∈ (0,0.001].

**S8 Table. IVW and MR-Egger estimates from the total PCa MVMR model adjusted for Lp(a) and AST**. Genetically elevated Lp(a) is significantly associated with total PCa through the IVW method. Associations of P<0.05 are shown in bold. .P ∈ (0.05,0.1], * P ∈ (0.01,0.05], ** P ∈ (0.001,0.01], *** P ∈ (0,0.001].

**S9 Table. IVW and MR-Egger estimates from the total PCa MVMR model adjusted for Lp(a) and creatinine**. Genetically elevated Lp(a) is significantly associated with total PCa through the IVW method and associated at borderline significance with total PCa with the MR-Egger approach. Associations of P<0.05 are shown in bold. .P ∈ (0.05,0.1], * P ∈ (0.01,0.05], ** P ∈ (0.001,0.01],

*** P ∈ (0,0.001].

**S10 Table. Association between SNPs and each lipid biomarker and BMI**. The following list includes IVs used in the main analysis, MVMR (adjusted for HDL,LDL,TG,Lp(a) and BMI) and sensitivity analyses 1-4.

**S11 Table. Association between SNPs and total PCa**. The following list includes IVs used in the main analysis, MVMR (adjusted for HDL,LDL,TG,Lp(a) and BMI) and sensitivity analyses 1-4. **S12 Table. Association between SNPs and advanced PCa**. The following list includes IVs used in the main analysis, MVMR (adjusted for HDL,LDL,TG,Lp(a) and BMI) and sensitivity analyses 1-4.

**S13 Table. Association between SNPs and early age onset PCa**. The following SNPs include IVs used in the main analysis, MVMR (adjusted for HDL,LDL,TG,Lp(a) and BMI) and sensitivity analyses 1-4.

**S14 Table. Association between SNPs, Lp(a), AST and total PCa**. The following SNPs include IVs used in the additional MVMR analysis on Lp(a), AST and total PCa only.

**S15 Table. Association between SNPs, Lp(a), creatinine and total PCa**. The following SNPs include IVs used in the additional MVMR analysis on Lp(a), creatinine and total PCa only.

**S1 File. Members from the PRACTICAL Consortium, CRUK, BPC3, CAPS and PEGASUS**

## References

1. Rawla P. Epidemiology of Prostate Cancer. World J Oncol. 2019;10(2):63–89.

2. Cancer R. Global cancer observatory [Internet]. [cited 2020 Jun 26]. Available from: https://gco.iarc.fr/

3. Loda M, Mucci LA, Mittelstadt ML, Van Hemelrijck M, Cotter MB. Pathology and epidemiology of cancer. In: Pathology and Epidemiology of Cancer. 2016. p. 1–670.

4. Pernar CH, Ebot EM, Wilson KM, Mucci LA. The Epidemiology of Prostate Cancer. Cold Spring Harb Perspect Med. 2018;8(12).

5. Tse LA, Ho WM, Wang F, He YH, Ng CF. Environmental risk factors of prostate cancer: a case-control study. Hong Kong Med J = Xianggang yi xue za zhi. 2018;24(4):30–3.

6. Moyad MA. Preventing aggressive prostate cancer with proven cardiovascular disease preventive methods. Asian J Androl. 2015;17(6):874–7.

7. Hurwitz LM, Agalliu I, Albanes D, Barry KH, Berndt SI, et al. Recommended Definitions of Aggressive Prostate Cancer for Etiologic Epidemiologic Research. JNCI J Natl Cancer Inst. 2021;113(6):727–34.

8. Katzke VA, Sookthai D, Johnson T, Kühn T, Kaaks R. Blood lipids and lipoproteins in relation to incidence and mortality risks for CVD and cancer in the prospective EPIC-Heidelberg cohort. BMC Med. 2017;15(1).

9. Jamnagerwalla J, Howard LE, Allott EH, Vidal AC, Moreira DM, Castro-Santamaria R, et al. Serum cholesterol and risk of high-grade prostate cancer: Results from the REDUCE study. Prostate Cancer Prostatic Dis. 2018;21(2):252–9.

10. Yu Peng L, Yu Xue Z, Fei LP, Cheng C, Ya Shuang Z, Da Peng L, et al. Cholesterol levels in blood and the risk of prostate cancer: A meta-analysis of 14 prospective studies. Cancer Epidemiol Biomarkers Prev. 2015;24(7):1086–93.

11. Bansal D, Undela K, D’Cruz S, Schifano F. Statin Use and Risk of Prostate Cancer: A Meta-Analysis of Observational Studies. PLoS One. 2012;7(10).

12. Tan P, Wei S, Tang Z, Gao L, Zhang C, Nie P, et al. LDL-lowering therapy and the risk of prostate cancer: A meta-analysis of 6 randomized controlled trials and 36 observational studies. Sci Rep. 2016;6.

13. Yarmolinsky J, Wade KH, Richmond RC, Langdon RJ, Bull CJ, Tilling KM, et al. Causal inference in cancer epidemiology: What is the role of mendelian randomization? Cancer Epidemiol Biomarkers Prev. 2018;27(9):995–1010.

14. Davies NM, Holmes M V., Davey Smith G. Reading Mendelian randomisation studies: A guide, glossary, and checklist for clinicians. BMJ. 2018;362.

15. Zheng J, Baird D, Borges M-C, Bowden J, Hemani G, Haycock P, et al. Recent Developments in Mendelian Randomization Studies. Curr Epidemiol Reports. 2017;4(4):330–45.

16. Bull CJ, Bonilla C, Holly JMP, Perks CM, Davies N, Haycock P, et al. Blood lipids and prostate cancer: a Mendelian randomization analysis. Cancer Med. 2016;5(6):1125–36.

17. Adams CD, Richmond R, Santos Ferreira DL, Spiller W, Tan V, Zheng J, et al. Circulating metabolic biomarkers of screen-detected prostate cancer in the ProtecT study. Cancer Epidemiol Biomarkers Prev. 2019;28(1):208–16.

18. Dominiczak MH, Caslake MJ. Apolipoproteins: Metabolic role and clinical biochemistry applications. Ann Clin Biochem. 2011;48(6):498–515.

19. Lab N. GWAS of UK Biobank biomarker measurements - Neale lab [Internet]. 2020 [cited 2020 Aug 16]. Available from: http://www.nealelab.is/blog/2019/9/16/biomarkers-gwas-results

20. Sinnott-Armstrong N, Tanigawa Y, Amar D, Mars N, Benner C, Aguirre M, et al. Genetics of 35 blood and urine biomarkers in the UK Biobank. Nat Genet. 2021;

21. Schumacher FR, Al Olama AA, Berndt SI, Benlloch S, Ahmed M, Saunders EJ, et al. Association analyses of more than 140,000 men identify 63 new prostate cancer susceptibility loci. Nat Genet. 2018;50(7):928–36.

22. Labrecque J, Swanson SA. Understanding the Assumptions Underlying Instrumental Variable Analyses: a Brief Review of Falsification Strategies and Related Tools. Curr Epidemiol Reports. 2018;5(3):214–20.

23. Rees JMB, Wood AM, Dudbridge F, Burgess S. Robust methods in Mendelian randomization via penalization of heterogeneous causal estimates. PLoS One. 2019;14(9).

24. Burgess S, Butterworth A TS. Mendelian randomization analysis with multiple genetic variants using summarized data. Genet Epidemiol. 2013;37(7):658–65.

25. Bowden J, Fabiola Del Greco M, Minelli C, Smith GD, Sheehan NA, Thompson JR. Assessing the suitability of summary data for two-sample mendelian randomization analyses using MR-Egger regression: The role of the I 2 statistic. Int J Epidemiol. 2016;45(6):1961– 74.

26. Rees JMB, Wood AM, Burgess S. Extending the MR-Egger method for multivariable Mendelian randomization to correct for both measured and unmeasured pleiotropy. Stat Med. 2017;36(29):4705–18.

27. Burgess S, Ference BA, Staley JR, Freitag DF, Mason AM, Nielsen SF, et al. Association of LPA variants with risk of coronary disease and the implications for lipoprotein(a)-lowering therapies: A mendelian randomization analysis. JAMA Cardiol. 2018;3(7):619–27.

28. Kronenberg F, Utermann G. Lipoprotein(a): Resurrected by genetics. J Intern Med. 2013;273(1):6–30.

29. Verbanck M, Chen CY, Neale B, Do R. Detection of widespread horizontal pleiotropy in causal relationships inferred from Mendelian randomization between complex traits and diseases. Nat Genet. 2018;50(5):693–8.

30. Burgess S, Foley CN, Allara E, Staley JR, Howson JMM. A robust and efficient method for Mendelian randomization with hundreds of genetic variants. Nat Commun. 2020;11(1).

31. Maranhão RC, Carvalho PO, Strunz CC, Pileggi F. Lipoprotein (a): Structure, pathophysiology and clinical implications. Arq Bras Cardiol. 2014;103(1):76–84.

32. Wang A, Lazo M, Ballentine Carter H, Groopman JD, Nelson WG, Platz EA. Association between Liver Fibrosis and Serum PSA among U.S. Men: National Health and Nutrition Examination Survey (NHANES), 2001-2010. Cancer Epidemiol Biomarkers Prev. 2019;28(8):1331–8.

33. Bañez LL, Loftis RM, Freedland SJ, Presti JC, Aronson WJ, Amling CL, et al. The influence of hepatic function on prostate cancer outcomes after radical prostatectomy. Prostate Cancer Prostatic Dis. 2010;13(2):173–7.

34. Hopewell JC, Haynes R, Baigent C. The role of lipoprotein (a) in chronic kidney disease. J Lipid Res. 2018;59(4):577–85.

35. Weinstein SJ, Mackrain K, Stolzenberg-Solomon RZ, Selhub J, Virtamo J, Albanes D. Serum creatinine and prostate cancer risk in a prospective study. Cancer Epidemiol Biomarkers Prev. 2009;18(10):2643–9.

36. Schmidt K, Noureen A, Kronenberg F, Utermann G. Structure, function, and genetics of lipoprotein (a). J Lipid Res. 2016;57(8):1339–59.

37. Van Capelleveen JC, Van Der Valk FM, Stroes ESG. Current therapies for lowering lipoprotein (a). J Lipid Res. 2016;57(9):1612–8.

38. Borgquist S, Butt T, Almgren P, Shiffman D, Stocks T, Orho-Melander M, et al. Apolipoproteins, lipids and risk of cancer. Int J Cancer. 2016;138(11):2648–56.

39. Wang FM, Zhang Y. High Lipoprotein(a) Level Is Independently Associated with Adverse Clinicopathological Features in Patients with Prostate Cancer. Dis Markers. 2019;2019.

40. Perez-Cornago A, Fensom GK, Andrews C, Watts EL, Allen NE, Martin RM, et al. Examination of potential novel biochemical factors in relation to prostate cancer incidence and mortality in UK Biobank. Br J Cancer. 2020;123(12):1808–17.

41. Murtola TJ, Syvälä H, Pennanen P, Bläuer M, Solakivi T, Ylikomi T, et al. The importance of LDL and Cholesterol metabolism for prostate epithelial cell growth. PLoS One. 2012;7(6).

42. Boffa MB, Koschinsky ML. Oxidized phospholipids as a unifying theory for lipoprotein(a) and cardiovascular disease. Nat Rev Cardiol. 2019;16(5):305–18.

43. Enkhmaa B, Anuurad E, Berglund L. Lipoprotein (a): Impact by ethnicity and environmental and medical conditions. J Lipid Res. 2016;57(7):1111–25.

44. Rebbeck TR. Prostate cancer disparities by race and ethnicity: From nucleotide to neighborhood. Cold Spring Harb Perspect Med. 2018;8(9).

45. Thomas JK, Mir H, Kapur N, Singh S. Racial diLerences in immunological landscape modifiers contributing to disparity in prostate cancer. Cancers (Basel). 2019;11(12).

46. Tang W, Wallace TA, Yi M, Magi-Galluzzi C, Dorsey TH, Onabajo OO, et al. IFNL4-ΔG allele is associated with an interferon signature in tumors and survival of African-American men with prostate cancer. Clin Cancer Res. 2018;24(21):5471–81.

47. Orsó E, Schmitz G. Lipoprotein(a) and its role in inflammation, atherosclerosis and malignancies. Clin Res Cardiol Suppl. 2017;12:31–7.

48. Fatumo S. The opportunity in African genome resource for precision medicine. EBioMedicine. 2020;54.

49. Haycock PC, Burgess S, Wade KH, Bowden J, Relton C, Smith GD. Best (but oft-forgotten) practices: The design, analysis, and interpretation of Mendelian randomization studies. Am J Clin Nutr. 2016;103(4):965–78.

50. Platz EA, Till C, Goodman PJ, Parnes HL, Figg WD, Albanes D, et al. Men with low serum cholesterol have a lower risk of high-grade prostate cancer in the placebo arm of the prostate cancer prevention trial. Cancer Epidemiol Biomarkers Prev. 2009;18(11):2807–13.

51. Platz EA, Clinton SK, Giovannucci E. Association between plasma cholesterol and prostate cancer in the PSA era. Int J Cancer. 2008;123(7):1693–8.

52. Burgess S, Butterworth A, Malarstig A, Thompson SG. Use of Mendelian randomisation to assess potential benefit of clinical intervention. BMJ. 2012;345.

53. Zuber V, Colijn JM, Klaver C, Burgess S. Selecting causal risk factors from high-throughput experiments using multivariable mendelian randomization. bioRxiv. 2018;

54. Kronenberg F. Human Genetics and the Causal Role of Lipoprotein(a) for Various Diseases. Cardiovasc Drugs Ther. 2016;30(1):87–100.

55. Viney NJ, van Capelleveen JC, Geary RS, Xia S, Tami JA, Yu RZ, et al. Antisense oligonucleotides targeting apolipoprotein(a) in people with raised lipoprotein(a): two randomised, double-blind, placebo-controlled, dose-ranging trials. Lancet. 2016;388(10057):2239–53.

