## Supplementary Tables 1-15 and S1 File for "The relationship between Lipoprotein A and other lipids with prostate cancer risk: A multivariable Mendelian randomisation study": S1 File.pdf

**PIs from the PRACTICAL (http://practical.icr.ac.uk/), CRUK, BPC3, CAPS, PEGASUS consortia:**

Rosalind A. Eeles[1,2], Christopher A. Haiman[3], Zsofia Kote-Jarai[1], Fredrick R. Schumacher[4,5], Sara Benlloch[6,1], Ali Amin Al Olama[6,7], Kenneth R. Muir[8], Sonja I. Berndt[9], David V. Conti[3], Fredrik Wiklund[10], Stephen Chanock[9], Ying Wang[11], Catherine M. Tangen[12], Jyotsna Batra[13,14], Judith A. Clements[13,14], APCB BioResource (Australian Prostate Cancer BioResource)[15,14], Henrik Grönberg[10], Nora Pashayan[16,17], Johanna Schleutker[18,19], Demetrius Albanes[9], Stephanie Weinstein[9], Alicja Wolk[20], Catharine M. L. West[21], Lorelei A. Mucci[22], Géraldine Cancel-Tassin[23,24], Stella Koutros[9], Karina Dalsgaard Sørensen[25,26], Eli Marie Grindedal[27], David E. Neal[28,29,30], Freddie C. Hamdy[31,32], Jenny L. Donovan[33], Ruth C. Travis[34], Robert J. Hamilton[35,36], Sue Ann Ingles[37], Barry S. Rosenstein[38], Yong-Jie Lu[39], Graham G. Giles[40,41,42], Robert J. MacInnis[40,41], Adam S. Kibel[43], Ana Vega[44,45,46], Manolis Kogevinas[47,48,49,50], Kathryn L. Penney[51], Jong Y. Park[52], Janet L. Stanford[53,54], Cezary Cybulski[55], Børge G. Nordestgaard[56,57], Sune F. Nielsen[56,57], Hermann Brenner[58,59,60], Christiane Maier[61], Jeri Kim[62], Esther M. John[63], Manuel R. Teixeira[64,65,66], Susan L. Neuhausen[67], Kim De Ruyck[68], Azad Razack[69], Lisa F. Newcomb[53,70], Davor Lessel[71], Radka Kaneva[72], Nawaid Usmani[73,74], Frank Claessens[75], Paul A. Townsend[76,77], Jose Esteban Castelao[78], Monique J. Roobol[79], Florence Menegaux[80], Kay-Tee Khaw[81], Lisa Cannon-Albright[82,83], Hardev Pandha[77], Stephen N. Thibodeau[84], David J. Hunter[85], Peter Kraft[86], William J. Blot[87,88], Elio Riboli[89]

[1]The Institute of Cancer Research, London, SM2 5NG, UK
[2]Royal Marsden NHS Foundation Trust, London, SW3 6JJ, UK
[3]Center for Genetic Epidemiology, Department of Preventive Medicine, Keck School of Medicine, University of Southern California/Norris Comprehensive Cancer Center, Los Angeles, CA 90015, USA
[4]Department of Population and Quantitative Health Sciences, Case Western Reserve University, Cleveland, OH 44106-7219, USA
[5]Seidman Cancer Center, University Hospitals, Cleveland, OH 44106, USA.
[6]Centre for Cancer Genetic Epidemiology, Department of Public Health and Primary Care, University of Cambridge, Strangeways Research Laboratory, Cambridge CB1 8RN, UK
[7]University of Cambridge, Department of Clinical Neurosciences, Stroke Research Group, R3, Box 83, Cambridge Biomedical Campus, Cambridge CB2 0QQ, UK
[8]Division of Population Health, Health Services Research and Primary Care, University of Manchester, Oxford Road, Manchester, M13 9PL, UK
[9]Division of Cancer Epidemiology and Genetics, National Cancer Institute, NIH, Bethesda, Maryland, 20892, USA
[10]Department of Medical Epidemiology and Biostatistics, Karolinska Institute, SE-171 77 Stockholm, Sweden

[11]Department of Population Science, American Cancer Society, 250 Williams Street, Atlanta, GA 30303, USA
[12]SWOG Statistical Center, Fred Hutchinson Cancer Research Center, Seattle, WA 98109, USA
[13]Australian Prostate Cancer Research Centre-Qld, Institute of Health and Biomedical Innovation and School of Biomedical Sciences, Queensland University of Technology, Brisbane QLD 4059, Australia
[14]Translational Research Institute, Brisbane, Queensland 4102, Australia
[15]Australian Prostate Cancer Research Centre-Qld, Queensland University of Technology, Brisbane; Prostate Cancer Research Program, Monash University, Melbourne; Dame Roma Mitchell Cancer Centre, University of Adelaide, Adelaide; Chris O'Brien Lifehouse and
[16]Department of Applied Health Research, University College London, London, WC1E 7HB, UK
[17]Centre for Cancer Genetic Epidemiology, Department of Oncology, University of Cambridge, Strangeways Laboratory, Worts Causeway, Cambridge, CB1 8RN, UK
[18]Institute of Biomedicine, University of Turku, Finland
[19]Department of Medical Genetics, Genomics, Laboratory Division, Turku University Hospital, PO Box 52, 20521 Turku, Finland
[20]Department of Surgical Sciences, Uppsala University, 75185 Uppsala, Sweden
[21]Division of Cancer Sciences, University of Manchester, Manchester Academic Health Science Centre, Radiotherapy Related Research, The Christie Hospital NHS Foundation Trust, Manchester, M13 9PL UK
[22]Department of Epidemiology, Harvard T. H. Chan School of Public Health, Boston, MA 02115, USA
[23]CeRePP, Tenon Hospital, F-75020 Paris, France.
[24]Sorbonne Universite, GRC n°5 , AP-HP, Tenon Hospital, 4 rue de la Chine, F-75020 Paris, France
[25]Department of Molecular Medicine, Aarhus University Hospital, Palle Juul-Jensen Boulevard 99, 8200 Aarhus N, Denmark
[26]Department of Clinical Medicine, Aarhus University, DK-8200 Aarhus N
[27]Department of Medical Genetics, Oslo University Hospital, 0424 Oslo, Norway
[28]Nuffield Department of Surgical Sciences, University of Oxford, Room 6603, Level 6, John Radcliffe Hospital, Headley Way, Headington, Oxford, OX3 9DU, UK
[29]University of Cambridge, Department of Oncology, Box 279, Addenbrooke's Hospital, Hills Road, Cambridge CB2 0QQ, UK
[30]Cancer Research UK, Cambridge Research Institute, Li Ka Shing Centre, Cambridge, CB2 0RE, UK
[31]Nuffield Department of Surgical Sciences, University of Oxford, Oxford, OX1 2JD, UK
[32]Faculty of Medical Science, University of Oxford, John Radcliffe Hospital, Oxford, UK
[33]Population Health Sciences, Bristol Medical School, University of Bristol, BS8 2PS, UK

[34]Cancer Epidemiology Unit, Nuffield Department of Population Health, University of Oxford, Oxford, OX3 7LF, UK
[35]Dept. of Surgical Oncology, Princess Margaret Cancer Centre, Toronto ON M5G 2M9, Canada
[36]Dept. of Surgery (Urology), University of Toronto, Canada
[37]Department of Preventive Medicine, Keck School of Medicine, University of Southern California/Norris Comprehensive Cancer Center, Los Angeles, CA 90015, USA
[38]Department of Radiation Oncology and Department of Genetics and Genomic Sciences, Box 1236, Icahn School of Medicine at Mount Sinai, One Gustave L. Levy Place, New York, NY 10029, USA
[39]Centre for Cancer Biomarker and Biotherapeutics, Barts Cancer Institute, Queen Mary University of London, John Vane Science Centre, Charterhouse Square, London, EC1M 6BQ, UK
[40]Cancer Epidemiology Division, Cancer Council Victoria, 615 St Kilda Road, Melbourne, VIC 3004, Australia
[41]Centre for Epidemiology and Biostatistics, Melbourne School of Population and Global Health, The University of Melbourne, Grattan Street, Parkville, VIC 3010, Australia
[42]Precision Medicine, School of Clinical Sciences at Monash Health, Monash University, Clayton, Victoria 3168, Australia
[43]Division of Urologic Surgery, Brigham and Womens Hospital, 75 Francis Street, Boston, MA 02115, USA
[44]Fundación Pública Galega Medicina Xenómica, Santiago de Compostela, 15706, Spain.
[45]Instituto de Investigación Sanitaria de Santiago de Compostela, Santiago De Compostela, 15706, Spain.
[46]Centro de Investigación en Red de Enfermedades Raras (CIBERER), Spain
[47]ISGlobal, Barcelona, Spain
[48]IMIM (Hospital del Mar Medical Research Institute), Barcelona, Spain
[49]Universitat Pompeu Fabra (UPF), Barcelona, Spain
[50]CIBER Epidemiología y Salud Pública (CIBERESP), 28029 Madrid, Spain
[51]Channing Division of Network Medicine, Department of Medicine, Brigham and Women's Hospital/Harvard Medical School, Boston, MA 02115, USA
[52]Department of Cancer Epidemiology, Moffitt Cancer Center, 12902 Magnolia Drive, Tampa, FL 33612, USA
[53]Division of Public Health Sciences, Fred Hutchinson Cancer Research Center, Seattle, Washington, 98109-1024, USA
[54]Department of Epidemiology, School of Public Health, University of Washington, Seattle, Washington 98195, USA
[55]International Hereditary Cancer Center, Department of Genetics and Pathology, Pomeranian Medical University, 70-115 Szczecin, Poland
[56]Faculty of Health and Medical Sciences, University of Copenhagen, 2200 Copenhagen, Denmark

[57]Department of Clinical Biochemistry, Herlev and Gentofte Hospital, Copenhagen University Hospital, Herlev, 2200 Copenhagen, Denmark
[58]Division of Clinical Epidemiology and Aging Research, German Cancer Research Center (DKFZ), D-69120, Heidelberg, Germany
[59]German Cancer Consortium (DKTK), German Cancer Research Center (DKFZ), D-69120 Heidelberg, Germany
[60]Division of Preventive Oncology, German Cancer Research Center (DKFZ) and National Center for Tumor Diseases (NCT), Im Neuenheimer Feld 460, 69120 Heidelberg, Germany
[61]Humangenetik Tuebingen, Paul-Ehrlich-Str 23, D-72076 Tuebingen, Germany
[62]The University of Texas M. D. Anderson Cancer Center, Department of Genitourinary Medical Oncology, 1515 Holcombe Blvd., Houston, TX 77030, USA
[63]Departments of Epidemiology & Population Health and of Medicine, Division of Oncology, Stanford Cancer Institute, Stanford University School of Medicine, Stanford, CA 94304 USA
[64]Department of Genetics, Portuguese Oncology Institute of Porto (IPO-Porto), 4200-072 Porto, Portugal
[65]Biomedical Sciences Institute (ICBAS), University of Porto, 4050-313 Porto, Portugal
[66]Cancer Genetics Group, IPO-Porto Research Center (CI-IPOP), Portuguese Oncology Institute of Porto (IPO-Porto), 4200-072 Porto, Portugal
[67]Department of Population Sciences, Beckman Research Institute of the City of Hope, 1500 East Duarte Road, Duarte, CA 91010, 626-256-HOPE (4673)
[68]Ghent University, Faculty of Medicine and Health Sciences, Basic Medical Sciences, Proeftuinstraat 86, B-9000 Gent
[69]Department of Surgery, Faculty of Medicine, University of Malaya, 50603 Kuala Lumpur, Malaysia
[70]Department of Urology, University of Washington, 1959 NE Pacific Street, Box 356510, Seattle, WA 98195, USA
[71]Institute of Human Genetics, University Medical Center Hamburg-Eppendorf, D-20246 Hamburg, Germany
[72]Molecular Medicine Center, Department of Medical Chemistry and Biochemistry, Medical University of Sofia, Sofia, 2 Zdrave Str., 1431 Sofia, Bulgaria
[73]Department of Oncology, Cross Cancer Institute, University of Alberta, 11560 University Avenue, Edmonton, Alberta, Canada T6G 1Z2
[74]Division of Radiation Oncology, Cross Cancer Institute, 11560 University Avenue, Edmonton, Alberta, Canada T6G 1Z2
[75]Molecular Endocrinology Laboratory, Department of Cellular and Molecular Medicine, KU Leuven, BE-3000, Belgium
[76]Division of Cancer Sciences, Manchester Cancer Research Centre, Faculty of Biology, Medicine and Health, Manchester Academic Health Science Centre, NIHR Manchester Biomedical Research Centre, Health Innovation Manchester, Univeristy of Manchester, M13 9WL
[77]The University of Surrey, Guildford, Surrey, GU2 7XH, UK

[78]Genetic Oncology Unit, CHUVI Hospital, Complexo Hospitalario Universitario de Vigo, Instituto de Investigación Biomédica Galicia Sur (IISGS), 36204, Vigo (Pontevedra), Spain
[79]Department of Urology, Erasmus University Medical Center, 3015 CE Rotterdam, The Netherlands
[80]"Exposome and Heredity", CESP (UMR 1018), Faculté de Médecine, Université Paris-Saclay, Inserm, Gustave Roussy, Villejuif
[81]Clinical Gerontology Unit, University of Cambridge, Cambridge, CB2 2QQ, UK
[82]Division of Epidemiology, Department of Internal Medicine, University of Utah School of Medicine, Salt Lake City, Utah 84132, USA
[83]George E. Wahlen Department of Veterans Affairs Medical Center, Salt Lake City, Utah 84148, USA
[84]Department of Laboratory Medicine and Pathology, Mayo Clinic, Rochester, MN 55905, USA
[85]Nuffield Department of Population Health, University of Oxford, United Kingdom
[86]Program in Genetic Epidemiology and Statistical Genetics, Department of Epidemiology, Harvard School of Public Health, Boston, MA, USA
[87]Division of Epidemiology, Department of Medicine, Vanderbilt University Medical Center, 2525 West End Avenue, Suite 800, Nashville, TN 37232 USA.
[88]International Epidemiology Institute, Rockville, MD 20850, USA
[89]Department of Epidemiology and Biostatistics, School of Public Health, Imperial College London, SW7 2AZ, UK

CRUK and PRACTICAL consortium

This work was supported by the Canadian Institutes of Health Research, European Commission's Seventh Framework Programme grant agreement n° 223175 (HEALTH-F2-2009-223175), Cancer Research UK Grants C5047/A7357, C1287/A10118, C1287/A16563, C5047/A3354, C5047/A10692, C16913/A6135, and The National Institute of Health (NIH) Cancer Post-Cancer GWAS initiative grant: No. 1 U19 CA 148537-01 (the GAME-ON initiative).

We would also like to thank the following for funding support: The Institute of Cancer Research and The Everyman Campaign, The Prostate Cancer Research Foundation, Prostate Research Campaign UK (now PCUK), The Orchid Cancer Appeal, Rosetrees Trust, The National Cancer Research Network UK, The National Cancer Research Institute (NCRI) UK. We are grateful for support of NIHR funding to the NIHR Biomedical Research Centre at The Institute of Cancer Research, The Royal Marsden NHS Foundation Trust, and Manchester NIHR Biomedical Research Centre. The Prostate Cancer Program of Cancer Council Victoria also acknowledge grant support from The National Health and Medical Research Council, Australia (126402, 209057, 251533, , 396414, 450104, 504700, 504702, 504715, 623204, 940394, 614296,), VicHealth, Cancer Council Victoria, The Prostate Cancer Foundation of Australia, The Whitten Foundation, PricewaterhouseCoopers, and Tattersall's. EAO, DMK, and EMK acknowledge the Intramural Program of the National Human Genome Research Institute for their support.

Genotyping of the OncoArray was funded by the US National Institutes of Health (NIH) [U19 CA 148537 for ELucidating Loci Involved in Prostate cancer SuscEptibility (ELLIPSE) project and X01HG007492 to the Center for Inherited Disease Research (CIDR) under contract number HHSN268201200008I]. Additional analytic support was provided by NIH NCI U01 CA188392 (PI: Schumacher).

Research reported in this publication also received support from the National Cancer Institute of the National Institutes of Health under Award Numbers U10 CA37429 (CD Blanke), and UM1 CA182883 (CM Tangen/IM Thompson). The content is solely the responsibility of the authors and does not necessarily represent the official views of the National Institutes of Health.

Funding for the iCOGS infrastructure came from: the European Community's Seventh Framework Programme under grant agreement n° 223175 (HEALTH-F2-2009-223175) (COGS), Cancer Research UK (C1287/A10118, C1287/A 10710, C12292/A11174, C1281/A12014, C5047/A8384, C5047/A15007, C5047/A10692, C8197/A16565), the National Institutes of Health (CA128978) and Post-Cancer GWAS initiative (1U19 CA148537, 1U19 CA148065 and 1U19 CA148112 - the GAME-ON initiative), the Department of Defence (W81XWH-10-1-0341), the Canadian Institutes of Health Research (CIHR) for the CIHR Team in Familial Risks of Breast Cancer, Komen Foundation for the Cure, the Breast Cancer Research Foundation, and the Ovarian Cancer Research Fund.

BPC3

The BPC3 was supported by the U.S. National Institutes of Health, National Cancer Institute (cooperative agreements U01-CA98233 to D.J.H., U01-CA98710 to S.M.G., U01-CA98216 to E.R., and U01-CA98758 to B.E.H., and Intramural Research Program of NIH/National Cancer Institute, Division of Cancer Epidemiology and Genetics).

CAPS

CAPS GWAS study was supported by the Cancer Risk Prediction Center (CRisP; www.crispcenter.org), a Linneus Centre (Contract ID 70867902) financed by the Swedish Research Council, (grant no K2010-70X-20430-04-3), the Swedish Cancer Foundation (grant no 09-0677), the Hedlund Foundation, the Soederberg Foundation, the Enqvist Foundation, ALF funds from the Stockholm County Council. Stiftelsen Johanna Hagstrand och Sigfrid Linner's Minne, Karlsson's Fund for urological and surgical research.

PEGASUS

PEGASUS was supported by the Intramural Research Program, Division of Cancer Epidemiology and Genetics, National Cancer Institute, National Institutes of Health.
